# Coronary artery disease and prostate cancer share a common genetic risk mediated through Lipoprotein(a)

**DOI:** 10.1101/2022.11.08.22282072

**Authors:** Richard A. Baylis, Fudi Wang, Hua Gao, Caitlin Bell, Lingfeng Luo, Kevin T. Nead, Tomas G. Neilan, Elsie Gyang Ross, Derek Klarin, Nicholas J. Leeper

## Abstract

The genetics underlying cancer and cardiovascular disease have been studied for decades. However, despite sharing numerous risk factors and pathologic features, the contribution of shared genetics in these diseases remains poorly defined. Our study sought to identify common genetic factors between these two diseases. We found a unique relationship between coronary artery disease and prostate cancer that appears to be mediated through germline genetic variation in LPA.

## Main Text

The advent of unbiased, genome-wide single nucleotide polymorphism sequencing has dramatically advanced our understanding of the pathophysiology underlying the two leading causes of death – cancer and cardiovascular disease. It is now well appreciated that patients being treated for cancer are at heightened risk for cardiovascular disease,^1,2^ and the expanding field of cardio-oncology focuses on characterizing and managing this risk. It has also been recognized that cancer and coronary artery disease (CAD) share numerous overlapping risk factors,^3^ and that mitigation efforts meant to benefit CAD also appear to decrease incident cancer.^4^ In addition, epidemiologic studies have shown that patients with CAD have an increased incidence of cancer development even when accounting for shared risk factors.^5,6^ Despite significant progress in our understanding of the genetic regulators of these two diseases, we know remarkably little about the role of shared genetic factors that may predispose to both cancer and cardiovascular disease. Our central hypothesis is that the relationship between cancer and CAD is mediated, in part, through shared genetic predisposition, and that understanding how genetics influence their interaction could facilitate future diagnostic and treatment efforts.

To determine which, if any, cancers are genetically correlated with CAD, we performed linkage disequilibrium score regression (LDSC)^7^ using summary statistics from 17 common cancers in the UK Biobank (UKBB) and CAD from CARDIoGRAMplusC4D^8^ (**Figure 1A**). Interestingly, prostate cancer (PCa) was the only cancer that was nominally correlated with CAD (Rg = 0.08, p value = 0.04). To validate this initial observation and understand how a patient’s genetic predisposition for CAD impacts their risk of developing cancer, we generated a polygenic risk score (PRS) for CAD using the CARDIoGRAMplusC4D dataset from Nikpay et al^9^ which did not include the UKBB patients and therefore minimized sample overlap. We then calculated the genetic risk for CAD among all patients in the UKBB, divided the population into PRS quintiles, and calculated odds ratios for the three most correlated cancers from our LDSC analysis for each CAD risk quintile. The incidence of lung cancer and melanoma showed no correlation with the CAD PRS (**Supplemental Figure 2**). However, consistent with the LDSC analysis, we found a clear positive correlation with PCa, such that those patients with the highest CAD PRS scores (80–100^th^ percentile) had the highest risk for prostate cancer (OR = 1.08, 95% CI 1.04-1.11, p value = 0.03) when compared to the lowest quintile group (0–20^th^ percentile; **Figure 1B**), further suggesting a unique genetic relationship between CAD and PCa. To ensure that the analysis was not being confounded by the inclusion of women in a male-only cancer, we repeated the PRS analysis excluding female subjects in the UKBB and observed a highly consistent effect (**Supplemental Figure 3**).

**Figure 1:**
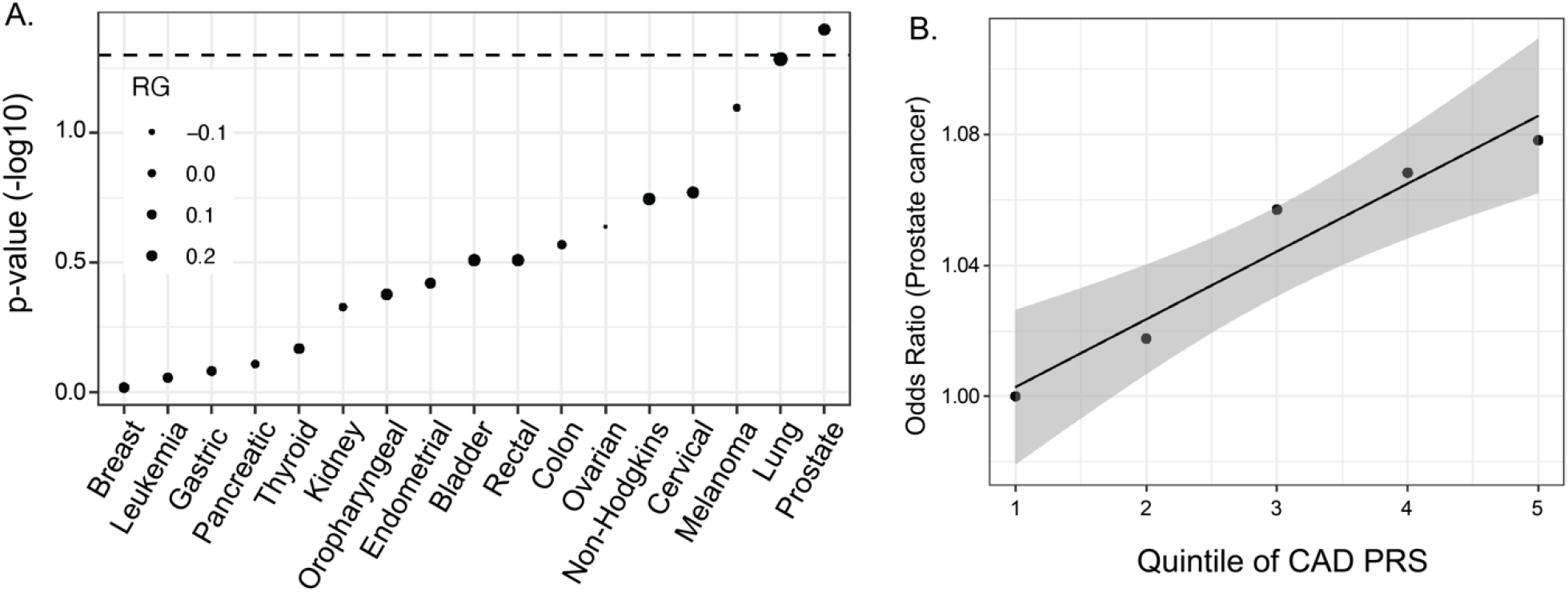
Genetic correlation between common cancers and CAD reveals a unique relationship between CAD and prostate cancer. **A**) Genetic correlation of GWAS from 17 common cancers in the UKBB with GWAS from coronary artery disease (CARDIoGRAMplusC4D) identifies prostate cancer as the only significantly correlated cancer type. **B**) Positive correlation of genetic risk for CAD through polygenic risk score quintiles with the prevalence of prostate cancer in the UKBB.

**Figure 2:**
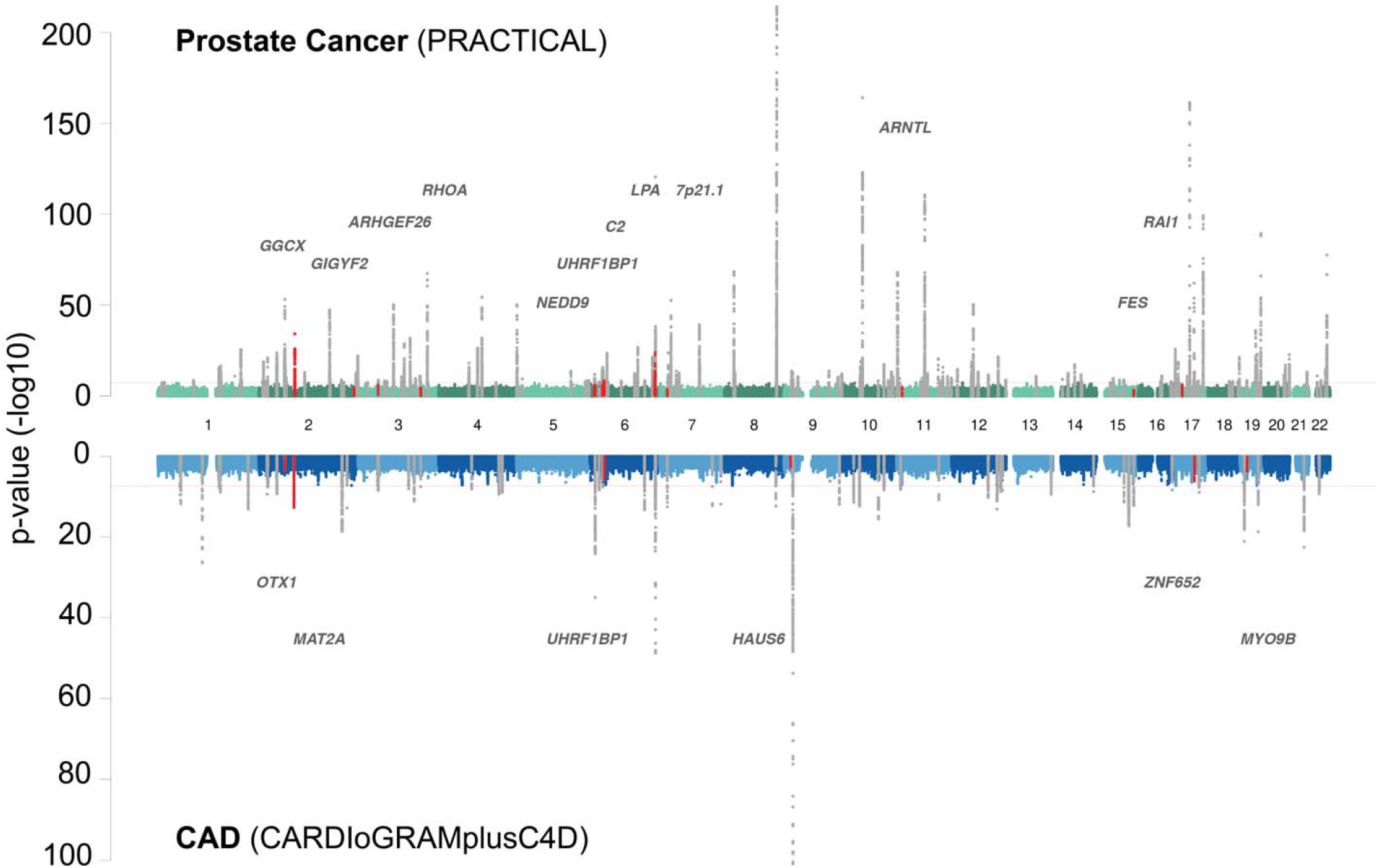
Identifying shared risk loci that achieve genome wide significance in one disease and nominal significance in the other. Miami plot comparing GWAS loci for prostate cancer (top of plot) and coronary artery disease (bottom of plot). The 18 loci denoted in red and annotated refer to GWS loci (i.e., p < 5×10^−8^) that are nominally significant (i.e., p< 0.005) in the opposite dataset. Specifically, the 12 loci in red on the upper Manhattan plot refer to GWS CAD loci that were found to be nominally significant in the prostate cancer dataset while on the lower Manhattan plot the 6 loci in red refer to GWS prostate cancer loci that are nominally significant in CAD.

**Figure 3.**
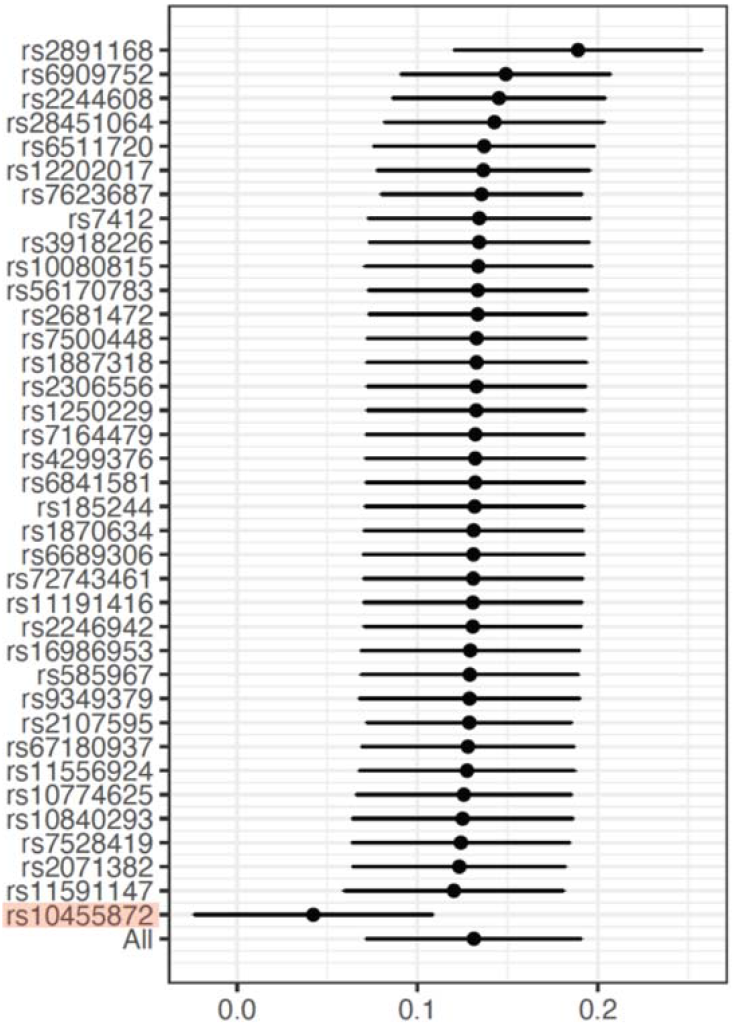
Determining the influence of each of the CAD loci used for the Mendelian Randomization reveals an outsized role of a single variant near LPA. Leave-one-out analysis of the Mendelian randomization using CAD as the exposure and prostate cancer as the outcome. Specifically, the Mendelian randomization was repeated with one of the CAD GWS loci removed from the instrumental variable. The large impact of removing rs10455872 from the analysis suggests that this loci has an outsized role in the association of CAD and prostate cancer.

To understand which loci were driving the genetic association of PCa and CAD, we determined if any genome-wide significant (GWS) loci for one disease demonstrated evidence of association in the other (p value < 0.005, **Figure 2**). To increase the statistical power of the analyses, we obtained a larger PCa-specific dataset from the PRACTICAL consortium. This analysis revealed 18 common loci between the two diseases – 6 GWS PCa loci with p < 0.005 in CAD and 12 GWS CAD loci with p < 0.005 in PCa (**Table 1**). Finally, to determine if there was a causal relationship between CAD genetics and PCa, we performed a Mendelian randomization analysis using all GWS (i.e., p value < 5×10^−8^) CAD loci^10^ as the instrumental variable using data from both UKBB and CARDIoGRAMplusC4D to maximize power.

**Table 1:** The 18 loci that were GWS in one dataset and at least nominally significant (p value < 0.005) in the opposite dataset.

| Prostate cancer loci that are GWS at <0.005 in CAD |  |  |  |  |  |  |
| --- | --- | --- | --- | --- | --- | --- |
| SNP ID | Gene | Chr | NEA | EA | OR | P value |
| rs58235267 | **OTX1** | 2 | C | G | 1.0290 | 1.0E-03 |
| rs2028900 | **MAT2A** | 2 | C | T | 1.0620 | 4.6E-13 |
| rs9469899 | **UHRF1BP1** | 6 | G | A | 1.0300 | 5.8E-04 |
| rs10122990 | **HAUS6** | 9 | A | C | 1.0260 | 2.5E-03 |
| rs2960158 | **ZNF652** | 17 | C | T | 1.0370 | 1.6E-04 |
| rs10412482 | **MYO9B** | 19 | C | T | 1.0290 | 2.4E-03 |

| CAD loci that are GWS at <0.005 in prostate cancer |  |  |  |  |  |  |
| --- | --- | --- | --- | --- | --- | --- |
| SNP ID | Gene | Chr | NEA | EA | OR | P value |
| rs7568458 | **GGCX** | 2 | T | A | 0.921 | 1.5E-36 |
| rs13003675 | **GIGYF2** | 2 | C | T | 0.978 | 1.3E-03 |
| rs12493885 | **ARHGEF26** | 3 | G | C | 1.047 | 1.9E-05 |
| rs7623687 | **RHOA** | 3 | C | A | 1.030 | 2.4E-03 |
| rs10455872 | **LPA** | 6 | G | A | 0.939 | 1.7E-05 |
| rs3130683 | **C2** | 6 | C | T | 0.965 | 5.6E-04 |
| rs4472337 | **UHRF1BP1** | 6 | C | T | 1.027 | 2.7E-03 |
| rs742115 | **NEDD9** | 6 | C | T | 1.023 | 2.2E-03 |
| rs2107595 | **7p21.1** | 7 | G | A | 0.967 | 7.3E-05 |
| rs3993105 | **ARNTL** | 11 | C | T | 1.022 | 2.7E-03 |
| rs2071382 | **FES** | 15 | C | T | 0.979 | 2.5E-03 |
| rs9897596 | **RAI1** | 17 | C | T | 1.027 | 6.8E-05 |

Interestingly, this analysis suggested that a genetically conferred increased risk of CAD was causally associated with an increased risk of PCa (beta 0.13, SE 0.05, p value = 0.03). To assess for particularly influential variants, we performed a leave-one-out analysis on the SNPs incorporated in the CAD genetic instrument. This demonstrated that most of the observed association was driven by a single variant (rs10455872) located in the LPA gene, a variant known to explain a substantial proportion of variation in plasma Lp(a) levels (**Figure 3**).^11,12^ Importantly, this SNP was also found among the GWS CAD loci that were nominally significant in the PRACTICAL consortium GWA data, suggesting that the positive MR result likely reflects independent pleiotropic effects of LPA on both CAD and PCa and may not support a causal association of CAD and PCa. This provides evidence of shared genetic risk between CAD and PCa that is primarily mediated through germline genetic variation in LPA.

## Discussion

Our study identifies a unique genetic relationship between PCa and CAD, which appears to be driven by pleiotropic effects of lipoprotein(a) on both diseases. To arrive at this conclusion, we screened 17 common cancers for genetic correlation with CAD, identifying PCa as the sole malignancy that showed genetic correlation. We then extended this finding by using an orthogonal strategy showing a positive correlation between the polygenic risk for CAD and prevalent prostate cancer. To look for overlapping loci that could explain this result, we found 18 GWS SNPs for one disease that were nominally significant for the other disease. Finally, we performed a Mendelian randomization study, which showed evidence of an association between CAD and PCa development. However, this appeared to be largely driven by a SNP at LPA, suggesting that Lp(a) is a shared pathogenic driver for both diseases. Taken together, our results identify a genetic link between CAD and PCa, further strengthening the emerging notion of overlapping pathogenesis of cardiovascular disease and cancer.

Previous studies have shown a clear increase in cardiovascular disease among patients with many different cancers.^1,2^ This increase could be explained by a host of causes including the presence of shared risk factors (smoking, metabolic syndrome, aging, etc), the known cardiotoxic effects of certain cancer treatments, and/or the result of common dysregulated processes known to exacerbate both diseases (e.g., chronic inflammation or metabolic dysfunction). Our study asked if shared genetic factors meaningfully contribute to this association and allowed us to demonstrate a clear role for shared genetics in the case of PCa. Future analyses with enhanced population genetics approaches and larger cancer genome datasets will determine if other malignancies similarly share a common genetic basis with CAD.

Cardiovascular disease is a leading cause of death among patients with PCa.^13^ However, understanding the epidemiologic interaction of CAD and PCa has been challenging given: 1.) the poor specificity of prostate specific antigen (PSA) screening and, 2.) detection bias from increased cancer screening in patients with chronic medical conditions like CAD. These challenges were partially overcome by a post-hoc analysis^14^ of the REDUCE Trial, an RCT studying dutasteride for primary prevention of prostate cancer. At the time of enrollment, a detailed medical history was obtained from each of the 5,843 patients, including CAD history, smoking status, medication use, and comorbid conditions. All patients, regardless of PSA levels, then underwent transrectal biopsy at 2 and 4 years, the gold standard for PCa diagnosis. Multivariate adjustment for numerous covariates (including age, comorbidities, smoking, and medication use) showed that men with CAD were at increased risk of developing prostate cancer (OR 1.35; 95% CI 1.08-1.67; p-value 0.007), regardless of whether they had low-or high-grade disease. Further, a separate study found that when prostate biopsies were analyzed for evidence of atherosclerosis, those samples from patients with PCa were more likely to have local atherosclerosis (OR 2.28) compared to tumor-negative autopsy prostate samples, demonstrating co-prevalent disease even at the histopathological level.^15^

From a translational perspective, a recent study of >90,000 US veterans with PCa cancer showed a high burden of modifiable cardiovascular risk factors which had been left untreated. These included hypertension (21.3% not on anti-hypertensive), hypercholesterolemia (47.6% not on lipid lowering therapy), and hyperglycemia (8.1% untreated), suggesting that healthcare providers are not adequately managing established risk factors which may be disproportionately important in this patient population.^16^ Our findings of a shared genetic predisposition highlight the importance of aggressive cardiovascular risk reduction in individuals with prostate cancer and provide a rationale for increased vigilance for signs of PCa among patients with CAD.

Beyond these general risk optimization considerations, our findings also highlight the opportunity for tailored therapies that could provide mechanistically-driven benefits across both conditions. LPA encodes for a serine proteinase, which is the main protein component of Lp(a), a highly heritable low-density lipoprotein variant. Polymorphisms in LPA are strongly associated with cardiovascular risk. For example, through Mendelian randomization genetically predicted levels of Lp(a) have been shown to be causally linked to CAD, PAD, and AAA.^17^ More recently, studies have suggested a link between Lp(a) and PCa, as well. One retrospective cohort study of several hundred patients with pathology-confirmed PCa tested whether any lipid components were associated with more severe disease. Lp(a) was the only lipid component found to have a relationship with PCa and, remarkably, patients with elevated Lp(a) were more likely to have higher risk pathologic features, higher levels of PSA, and more advanced stages of disease.^18^ Interestingly, this association with Lp(a) appears to be unique to PCa and was not seen with other common cancers.^19^ Finally, a Mendelian randomization study investigating genetically-predicted lipid components and their relationship with PCa found that genetically-predicted Lp(a) was associated with a higher risk for PCa.^20^ Given that the same lipoprotein may be causally-related to both diseases, it is exciting to hypothesize that the promising Lp(a) therapeutics currently under investigation may be able to reduce both CAD and PCa incidence – two of the leading causes of morbidity in men.^21^

In conclusion, significant effort has been made to understand how cancer treatment strategies impact a patient’s risk for cardiovascular disease, but much less attention has been placed on understanding the fundamental genetic relationship between these diseases. In this study, we sought to take advantage of the large amount of genetic data that has been independently generated for both diseases to try to understand how genetic risk for one impacts the other. Surprisingly, we found that prostate cancer was uniquely genetically associated with CAD, which may have important implications for cancer screening and integrated treatment approaches which could target both diseases simultaneously.

## Data Availability

GWAS summary statistics for 17 common cancers were obtained from the UK biobank (https://github.com/Wittelab/pancancer_pleiotropy) and for CAD from meta-analysis of UK Biobank SOFT CAD GWAS with the CARDIoGRAMplusC4D 1000 genomes-based GWAS and the Myocardial Infarction Genetics and CARDIoGRAM Exome(http://www.cardiogramplusc4d.org/). When prostate cancer was identified as the only cancer that correlated with CAD, prostate cancer GWAS summary statistics were obtained from the PRACTICAL consortium to increase the power for further analyses (http://practical.icr.ac.uk/blog/).

## Funding

N.L. was supported by NIH R35 HL144475 and the American Heart Association EIA34770065.

R.A.B. was supported by NIH F30 HL136188

K.T.N. is a Cancer Prevention Research Institute of Texas (CPRIT) Scholars in Cancer Research and is supported by CPRIT RR190077 and NCI K08CA263313

## Author Contributions

R.A.B. and N.L. conceptualized the project. F.W. performed the analyses, generated figures, and contributed critical intellectual input. R.A.B., F.W., D.K., N.L. wrote the manuscript. H.G., C.B., L.L., K.T.N., T.G.N., E.R., J.E., provided critical intellectual input and revised the manuscript. N.L. and D.K. supervised the project and data analyses.

## Acknowledgements

We thank the PRACTICAL Consortium for providing access to the prostate cancer GWAS summary statistics.

---

**Supplemental Figure 1:**
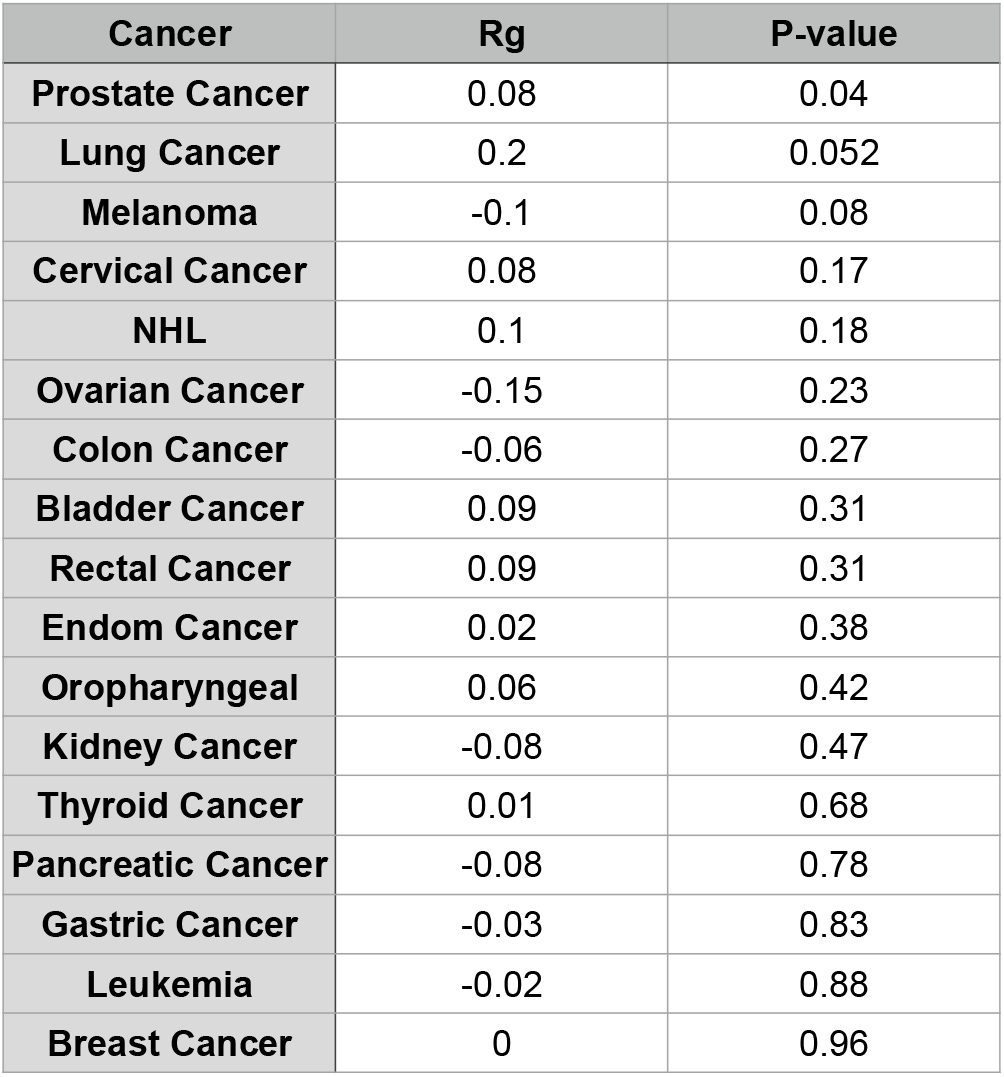
Table with each of the 17 cancers from the UKBB with their LDSC correlation score, Rg, and p-value, identifying prostate cancer as the only cancer with a significant signal.

**Supplemental Figure 2:**
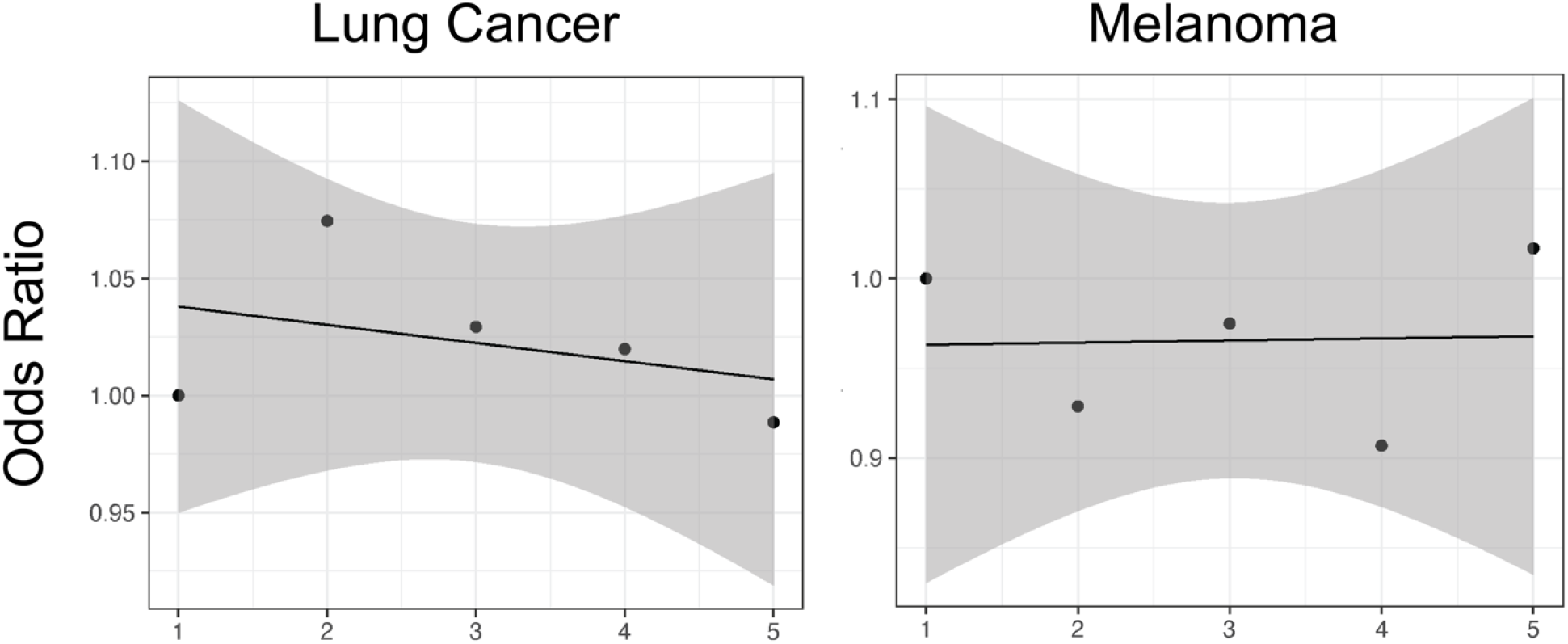
There is no clear correlation between CAD risk quintiles in the UKBB and prevalence of lung cancer or melanoma, suggesting the relationship of CAD and prostate cancer is unique.

**Supplemental Figure 3:**
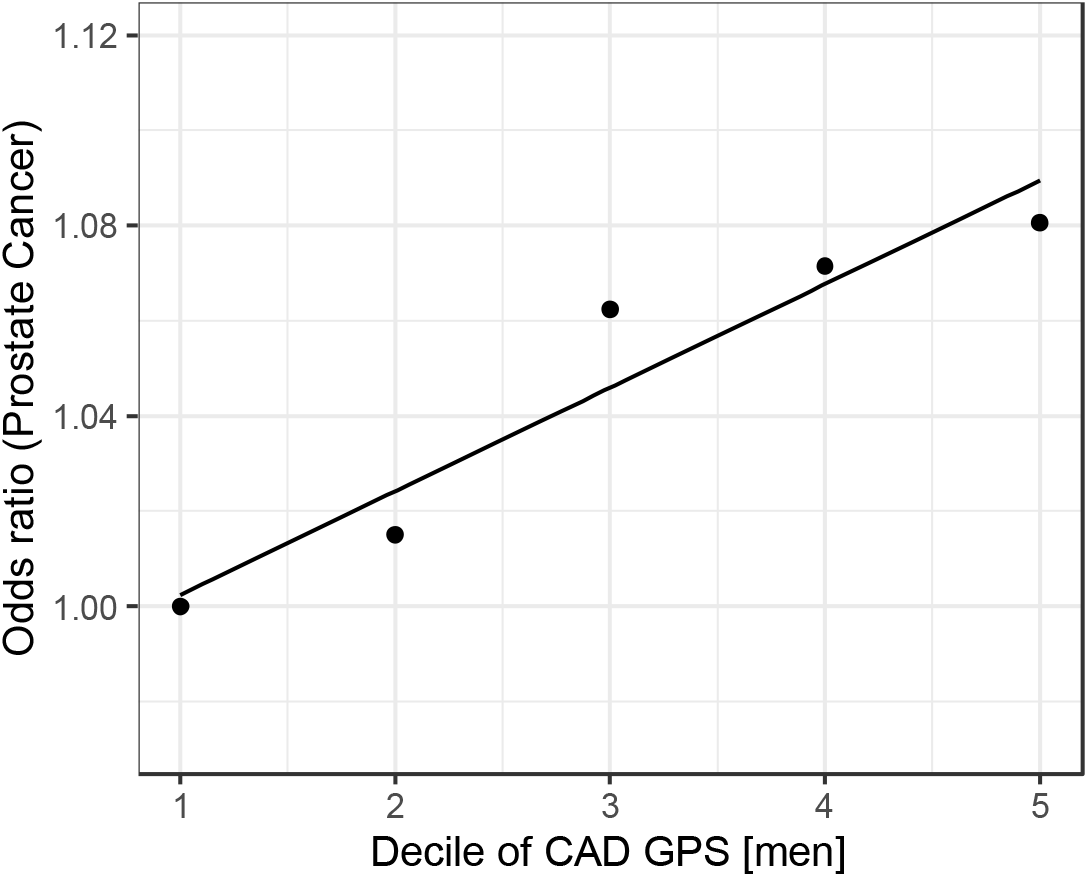
Replication of the PRS analysis using only male patients from the UKBB

## Notes

**Competing Interests:** The authors have declared that no conflict of interest exists.

### Competing Interest Statement

The authors have declared no competing interest.

## References

1. Armenian, S. H. et al. Cardiovascular Disease Among Survivors of Adult-Onset Cancer: A Community-Based Retrospective Cohort Study. JCO 34, 1122–1130 (2016).

2. Strongman, H. et al. Medium and long-term risks of specific cardiovascular diseases in survivors of 20 adult cancers: a population-based cohort study using multiple linked UK electronic health records databases. The Lancet 394, 1041–1054 (2019).

3. Handy, C. E. et al. Synergistic Opportunities in the Interplay Between Cancer Screening and Cardiovascular Disease Risk Assessment: Together We Are Stronger. Circulation 138, 727–734 (2018).

4. Rasmussen-Torvik, L. J. et al. Ideal Cardiovascular Health Is Inversely Associated With Incident Cancer: The Atherosclerosis Risk in Communities Study. Circulation 127, 1270–1275 (2013).

5. Suzuki, M. et al. Incidence of cancers in patients with atherosclerotic cardiovascular diseases. IJC Heart & Vasculature 17, 11–16 (2017).

6. Bell, C. F. et al. Risk of Cancer After Diagnosis of Cardiovascular Disease. http://medrxiv.org/lookup/doi/10.1101/2022.08.08.22278548 (2022) doi:10.1101/2022.08.08.22278548.

7. Gazal, S. et al. Linkage disequilibrium–dependent architecture of human complex traits shows action of negative selection. Nat Genet 49, 1421–1427 (2017).

8. the CARDIoGRAMplusC4D Consortium. A comprehensive 1000 Genomes–based genome-wide association meta-analysis of coronary artery disease. Nat Genet 47, 1121–1130 (2015).

9. Nikpay, M. et al. A comprehensive 1,000 Genomes-based genome-wide association meta-analysis of coronary artery disease. Nat Genet 47, 1121–1130 (2015).

10. Nelson, C. P. et al. Association analyses based on false discovery rate implicate new loci for coronary artery disease. Nat Genet 49, 1385–1391 (2017).

11. Clarke, R. et al. Genetic variants associated with Lp(a) lipoprotein level and coronary disease. N Engl J Med 361, 2518–2528 (2009).

12. Emdin, C. A. et al. Phenotypic Characterization of Genetically Lowered Human Lipoprotein(a) Levels. J Am Coll Cardiol 68, 2761–2772 (2016).

13. Shikanov, S., Kocherginsky, M., Shalhav, A. L. & Eggener, S. E. Cause-specific mortality following radical prostatectomy. Prostate Cancer Prostatic Dis 15, 106–110 (2012).

14. Thomas, J.-A. et al. Prostate Cancer Risk in Men with Baseline History of Coronary Artery Disease: Results from the REDUCE Study. Cancer Epidemiol Biomarkers Prev 21, 576–581 (2012).

15. Hager, M., Mikuz, G., Bartsch, G., Kolbitsch, C. & Moser, P. L. The association between local atherosclerosis and prostate cancer. BJU Int 99, 46–48 (2007).

16. Sun, L. et al. Assessment and Management of Cardiovascular Risk Factors Among US Veterans With Prostate Cancer. JAMA Netw Open 4, e210070 (2021).

17. Larsson, S. C. et al. Lipoprotein(a) in Alzheimer, Atherosclerotic, Cerebrovascular, Thrombotic, and Valvular Disease: Mendelian Randomization Investigation. Circulation 141, 1826–1828 (2020).

18. Wang, F.-M. & Zhang, Y. High Lipoprotein(a) Level Is Independently Associated with Adverse Clinicopathological Features in Patients with Prostate Cancer. Disease Markers 2019, 1–7 (2019).

19. Katzke, V. A., Sookthai, D., Johnson, T., Kühn, T. & Kaaks, R. Blood lipids and lipoproteins in relation to incidence and mortality risks for CVD and cancer in the prospective EPIC–Heidelberg cohort. BMC Med 15, 218 (2017).

20. Ioannidou, A. et al. The relationship between lipoprotein A and other lipids with prostate cancer risk: A multivariable Mendelian randomisation study. PLoS Med 19, e1003859 (2022).

21. Tsimikas, S. et al. Lipoprotein(a) Reduction in Persons with Cardiovascular Disease. N Engl J Med 382, 244–255 (2020).

