## Supplementary material for "Coronary artery disease and prostate cancer share a common genetic risk mediated through Lipoprotein(a)": Materials and Methods

**Study Materials**

GWAS summary statistics for 17 common cancers were obtained from the UK biobank (https://github.com/Wittelab/pancancer_pleiotropy) and for CAD from meta-analysis of UK Biobank SOFT CAD GWAS with the CARDIoGRAMplusC4D 1000 genomes-based GWAS and the Myocardial Infarction Genetics and CARDIoGRAM Exome^1^ (http://www.cardiogramplusc4d.org/). When prostate cancer was identified as the only cancer that correlated with CAD, prostate cancer GWAS summary statistics were obtained from the PRACTICAL consortium to increase the power for further analyses^2^ (http://practical.icr.ac.uk/blog/). Details on the study characteristic for these datasets have been described elsewhere^3^. Autosomal SNPs with a minor allele frequency (MAF) greater than 1% were included in the analyses as they were well imputed across studies, and sex chromosomes were excluded. R version 3.6.3 was used for statistical analyses.

**Genetic correlation between cancer and CAD**

LD score regression (LDSC, https:// github.com/bulik/ldsc) was performed using default parameters on GWAS summary statistics from the autosomal SNPs to calculate the genetic correlation between each cancer type and CAD. LDSC is a method for estimating genetic correlation based on selected functional annotations. In this study, we followed the analysis tutorial (https://github.com/bulik/ldsc/wiki/Heritability-and-Genetic-Correlation) using the recommended version 2.2 of the baseline-LD model with 97 annotations. Most samples from our study were of European ancestry, we performed the LDSC using European LD scores and allele frequencies from the 1000 Genomes Phase 3 project.

**Prostate cancer prevalence for each CAD-polygenic risk score quintile**

A polygenic risk score (PRS) for CAD was generated from CARDIoGRAMplusC4D (without UKBB data to minimize sample overlap)^4^ using the PRScs tool (https://github.com/getian107/PRScs). Briefly, PRScs applies a Bayesian shrinkage parameter to the effect size estimates for each SNP using GWAS summary statistics. The p-value threshold and the global shrinkage parameter, phi, were tuned to optimize the model’s performance, which was assessed using area under the curve (AUC) adjusting for sex, age, and the first five genetic principal components (PCs). The performance of each PRS was estimated using a non-overlapping set of subjects from the UK Biobank with 10-fold cross validation.

The best performing PRS score was used to evaluate the correlation of a CAD-specific PRS scores and prostate cancer prevalence across the population. To this end, the cohort was divided into quintiles based on each patient’s CAD-specific PRS value and the prevalence of prostate cancer within each quintile was determined.

**Mendelian randomization between CAD risk loci and Prostate Cancer**

Mendelian randomization (MR) analysis using the TwoSampleMR R package (version 0.4.22, https:// github.com/MRCIEU/TwoSampleMR) was used to performed with CAD as the exposure and prostate cancer (PRACTICAL) as the outcome. DNA sequence variants associated with CAD at genome wide significance (p < 5x10^-8^) with an r^2^<0.001 were selected as genetic instruments. All clumping and harmonization were completed using the TwoSampleMR package. Inverse-variance weighting (IVW) applying a multiplicative random effects model was used to study the effect of coronary artery disease risk loci on prostate cancer.

References:

1.	Nelson, C. P. *et al.* Association analyses based on false discovery rate implicate new loci for coronary artery disease. *Nat Genet* **49**, 1385–1391 (2017).

2.	Kote-Jarai, Z. *et al.* Multiple novel prostate cancer predisposition loci confirmed by an international study: the PRACTICAL Consortium. *Cancer Epidemiol Biomarkers Prev* **17**, 2052–2061 (2008).

3.	Rashkin, S. R. *et al.* Pan-cancer study detects genetic risk variants and shared genetic basis in two large cohorts. *Nat Commun* **11**, 4423 (2020).

4.	Nikpay, M. *et al.* A comprehensive 1,000 Genomes-based genome-wide association meta-analysis of coronary artery disease. *Nat Genet* **47**, 1121–1130 (2015).
